# AI detects a distributed blood metabolomic Systemotype associated with early stage ovarian cancer

**DOI:** 10.64898/2026.09.10.26362758

**Authors:** Hongyi Zhou, Jean-Luc Chaubard, Benedict Benigno, Jeffrey Skolnick

## Abstract

Early detection of ovarian cancer remains a clinical challenge because available blood biomarkers lack the sensitivity and specificity required for population screening. We tested whether early-stage ovarian cancer is associated with a distributed physiological state in the circulating metabolome. We analyzed untargeted metabolomic profiles from two independent retrospective cohorts: 91 serum samples (59 ovarian cancer, 32 healthy controls) and 83 plasma samples (63 ovarian cancer, 20 healthy controls). Assay-specific boosted decision trees were trained and evaluated independently within each cohort using five-fold cross-validation repeated over 50 randomized rounds. At selected operating points, mean cross-validated sensitivity and specificity were 99.0% and 99.8% in serum and 97.7% and 99.7% in plasma. Restricting inputs to strongly dysregulated features did not improve the overall sensitivity–false-positive-rate trade-off, and smaller panels reduced sensitivity. The cohorts shared 239 concordantly altered annotated features spanning lipid, amino-acid, steroid, central-carbon, and redox metabolism. These findings are consistent with a distributed metabolic response involving tumor and host, although tissue contributions were not measured. We propose that the classifier recognizes a metabolomic Systemotype, an integrated physiological state reflected in circulating metabolites. The results support further investigation of this framework.

## Introduction

Blood continuously exchanges metabolites, proteins, hormones, lipids, extracellular vesicles, and signaling molecules with tissues throughout the body. Its molecular composition therefore reflects integrated physiological activity. Clinical medicine exploits this property through circulating measurements that assess endocrine function ^1^, liver chemistry ^2^, and myocardial injury ^3^. These examples support a broader principle: blood provides a dynamic molecular readout of organismal physiology ^4,5^.

This perspective raises a fundamental question: does disease appear in blood primarily through a few disease-specific biomarkers, or through broader changes in organismal physiology? Most diagnostic strategies emphasize individual analytes or small panels whose concentrations differ between patients and controls. This approach has produced clinically useful assays. For many complex diseases, however, individual biomarkers have not achieved the sensitivity and specificity required for population screening, particularly when disease prevalence is low ^6,7^.

An alternative possibility is that disease is reflected in a distributed physiological state. Localized pathology can perturb metabolism, inflammatory signaling, endocrine regulation, vascular physiology, nutrient allocation, and immune function. Communication among tissues may propagate these perturbations, producing changes throughout the circulating metabolome. Individual differences may be modest, yet collectively they could define a reproducible physiological state ^8,9,10^.

We refer to such an integrated physiological state as a Systemotype: the condition of an organism arising from interactions among organs, tissues, cells, and molecular networks. A disease-associated Systemotype may reflect both pathological processes and adaptive host responses. The circulating metabolome is well suited to investigating this concept because metabolite concentrations reflect exchange among tissues and the balance of production, consumption, transport, modification, and clearance ^4,5,9^.

Artificial intelligence (AI) offers a means of recognizing this distributed organization. Machine-learning algorithms can integrate many variables whose joint information is difficult to capture in analyses of individual metabolites. In the framework proposed here, AI serves as an analytical tool for detecting a multivariate physiological state in the circulating metabolome ^11^.

Early-stage ovarian cancer (OC) provides a clinically relevant setting in which to investigate this hypothesis. OC remains a leading cause of gynecologic cancer mortality; many patients are diagnosed after dissemination, whereas outcomes are substantially better for stage I disease ^12^. Randomized screening trials illustrate the difficulty of translating earlier detection into a reduction in mortality ^6,7^. Because OC is uncommon, even a modest false-positive rate can generate many unnecessary investigations. CA125 is widely used for disease monitoring and evaluation of suspicious adnexal masses, but it lacks the performance required for population screening ^13^. Multimarker approaches have improved risk assessment in patients with pelvic masses; their performance in these populations does not establish suitability for general-population screening ^14,15,16^.

Previous studies have reported serum and plasma metabolomic or lipidomic signatures associated with OC, including early-stage disease ^17,18,19,20^. If early OC is associated with a reproducible blood Systemotype, recognizing that state could support diagnostic development and help explain the biological basis of metabolomic classification. Independent and prospective validation remains essential.

Here we address four questions. First, can AI distinguish early-stage OC from healthy controls using circulating metabolomic profiles? Second, does classification depend on a small set of strongly dysregulated features or on information distributed more broadly across the metabolome? Third, do independently analyzed serum and plasma cohorts share higher-order biochemical patterns? Finally, are these patterns consistent with physiological responses involving multiple tissues? The final question concerns biological interpretation; the present study does not directly test causal tissue contributions.

## Results

### Classification of ovarian cancer within independent serum and plasma cohorts

We analyzed untargeted metabolomic profiles from two independently processed cohorts (Figure 1; Methods). Batch 1 comprised 3,433 metabolite features from 91 serum samples (59 OC, 32 healthy controls), and Batch 2 comprised 3,625 features from 83 plasma samples (63 OC, 20 healthy controls). Features were partitioned into four assays: lipid_pos, lipid_neg, polar_pos, and polar_neg. Increased and decreased features are listed in the supplementary files sera_up.tsv, sera_down.tsv, plasma_up.tsv, and plasma_down.tsv. A separate boosted decision-tree model was trained for each assay ^21^. Assay-level calls were combined using an OR rule: a sample was classified as OC if any assay predicted OC. Within each cohort, five-fold cross-validation was repeated over 50 randomized rounds, yielding 250 held-out evaluations. The one million group-assignment swaps used to randomize each partition were not one million independent validation runs. The two cohorts were modeled and evaluated separately.

**Figure 1.**
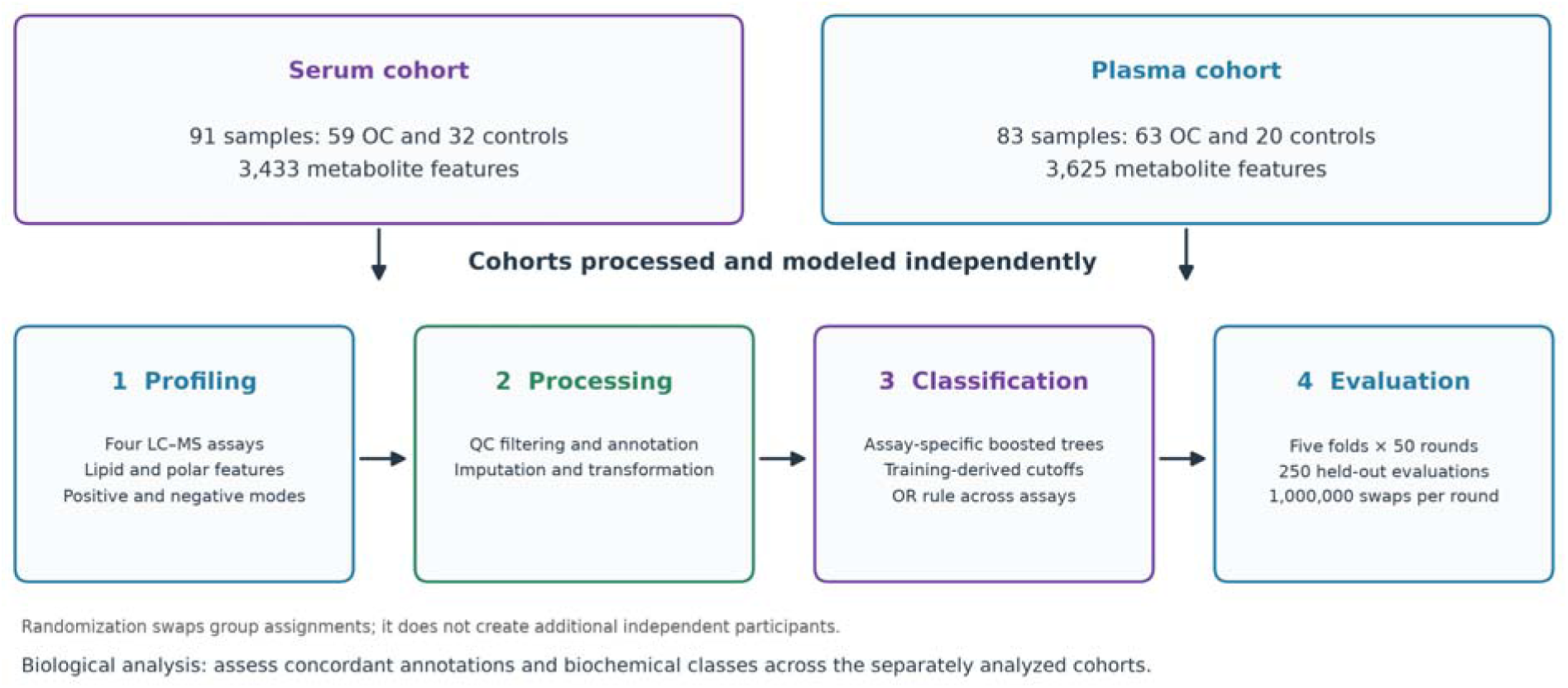
Analysis workflow. Serum and plasma cohorts were processed and modeled independently. The workflow comprised untargeted metabolomic profiling, feature processing, assay-specific boosted decision trees, repeated five-fold cross-validation, and biochemical class analysis ^21^. The plasma analysis provides replication in a separately modeled cohort; it is not an external test of a serum-trained model.

Using the full feature sets, the models achieved high mean cross-validated sensitivity at low false-positive rates (Table 1; Figure 2). Across the six reported parameter settings, sensitivity ranged from 90.3% to 99.1% and false-positive rates from 0.0% to 1.1%. At m = 5 and δ = 0.05, the reported mean false-positive rate was 0.0%, with sensitivities of 95.5% in serum and 90.3% in plasma. Among operating points with mean specificity above 99.5%, serum sensitivity was 99.0% at 99.8% specificity (m = 5, δ = 0.001), and plasma sensitivity was 97.7% at 99.7% specificity (m = 5, δ = 0.01). These retrospective estimates require confirmation in independent prospective cohorts. A reported mean false-positive rate of 0.0% does not establish perfect specificity in a screening population.

**Table 1.** Mean sensitivity and false-positive rate using the full feature sets. Values summarize five-fold cross-validation repeated over 50 randomized rounds (250 held-out evaluations per cohort).

| Cutoff parameters<sup>a</sup> | Serum sensitivity | Serum false-positive rate<sup>b</sup> | Plasma sensitivity | Plasma false-positive rate<sup>b</sup> |
| --- | --- | --- | --- | --- |
| m = 2, $\delta$ = 0.05 | 99.1% | 0.9% | 97.8% | 1.1% |
| m = 3, $\delta$ = 0.05 | 98.2% | 0.4% | 97.1% | 0.7% |
| m = 4, $\delta$ = 0.05 | 96.9% | 0.2% | 94.4% | 0.2% |
| **m = 5, $\delta$ = 0.05** | **95.5%** | **0.0%** | **90.3%** | **0.0%** |
| m = 5, $\delta$ = 0.01 | 98.6% | 0.13% | 97.7% | 0.3% |
| m = 5, $\delta$ = 0.001 | 99.0% | 0.2% | 97.9% | 0.6% |

a Cutoff parameters m and $\delta$ are defined in Methods, Equation 2. The parameter m specifies the ranking gap, and $\delta$ is an offset added to the selected score. Bold text marks the parameter setting used for the matched comparisons of feature-set size.
b Specificity (%) = 100 – false-positive rate (%). All performance tables report means rounded to the precision supplied; 0.0% should not be interpreted as a population-level guarantee of zero false positives.

**Figure 2.**
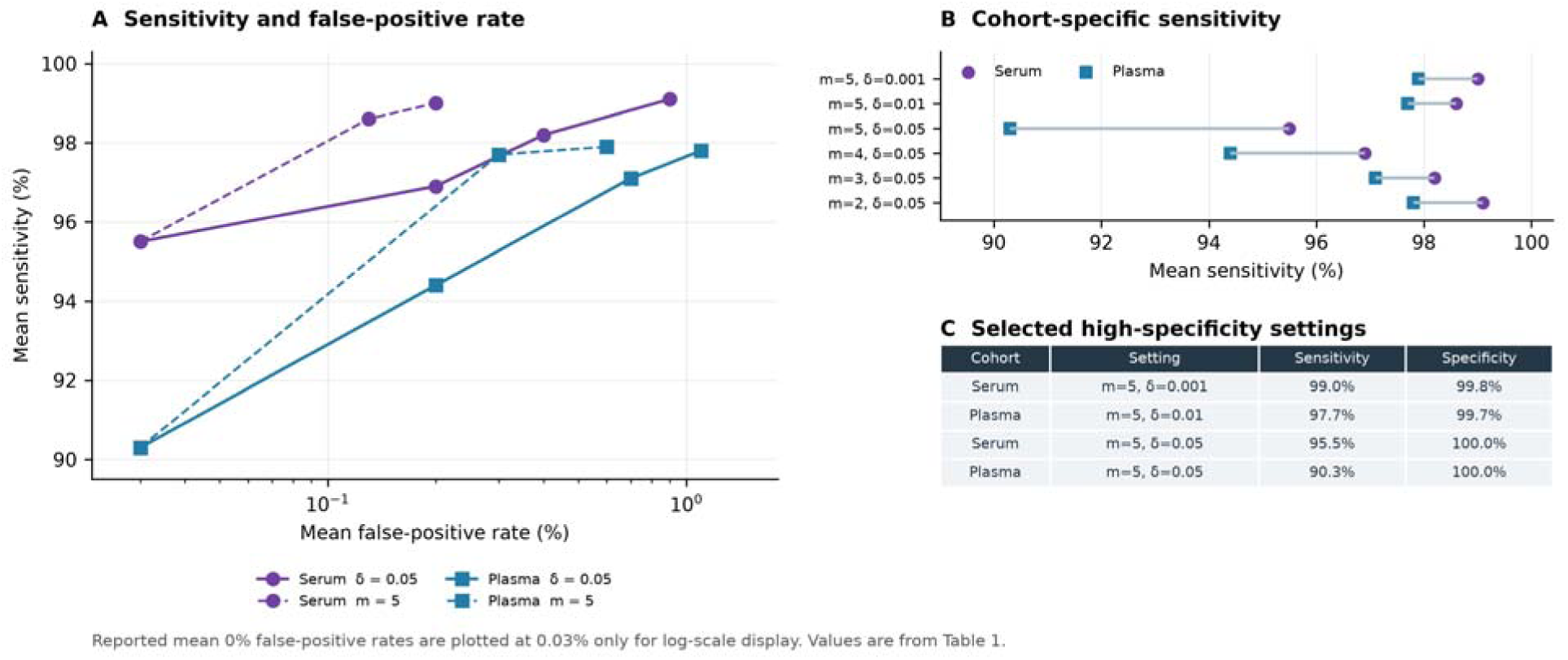
Cross-validated classification performance at the reported decision thresholds. The parameter m specifies the ranking gap used to select a training-derived score, and δ is an additive offset (Methods, Equation 2). (A) Sensitivity versus false-positive rate using the full feature sets in the independently modeled serum and plasma cohorts. (B) Sensitivity at the six reported parameter settings. (C) High-specificity operating points in Table 1. Mean false-positive rates reported as 0% are plotted at 0.03% in panel A to permit display on a logarithmic axis. These curves summarize within-cohort cross-validation rather than external validation.

### Restricting features to strongly dysregulated metabolites

To assess whether classification depended primarily on strongly dysregulated features, we restricted the inputs to features meeting the Mann–Whitney U test criterion |z| > 2 ^22^. This reduced the feature count from 3,433 to 2,160 in serum and from 3,625 to 2,037 in plasma. Relative to the full feature sets, this restriction did not improve the overall sensitivity–false-positive-rate trade-off (Tables 1 and 2). False-positive rates were higher at nearly every operating point; the exception was plasma at m = 2 and δ = 0.05, where the rate decreased from 1.1% to 0.9%. Sensitivity changed modestly and did not improve consistently. We also evaluated smaller panels selected from the most strongly increased and decreased features ranked by z-score (Tables 3A–C).

**Table 2.** Mean cross-validated sensitivity and false-positive rate using features meeting the Mann–Whitney criterion |z| > 2. Cutoff definitions and evaluation rounds are as in Table 1.

| Cutoff parameters | Serum sensitivity | Serum false-positive rate | Plasma sensitivity | Plasma false-positive rate |
| --- | --- | --- | --- | --- |
| $m = 2, \delta = 0.05$ | 98.9% | 1.4% | 97.9% | 0.9% |
| $m = 3, \delta = 0.05$ | 97.9% | 0.7% | 96.3% | 0.9% |
| $m = 4, \delta = 0.05$ | 96.9% | 0.3% | 93.5% | 0.3% |
| **$m = 5, \delta = 0.05$** | **95.2%** | **0.06%** | **88.8%** | **0.1%** |
| $m = 5, \delta = 0.01$ | 98.8% | 0.4% | 97.8% | 0.6% |
| $m = 5, \delta = 0.001$ | 99.1% | 0.5% | 98.1% | 0.7% |

**Table 3A.**
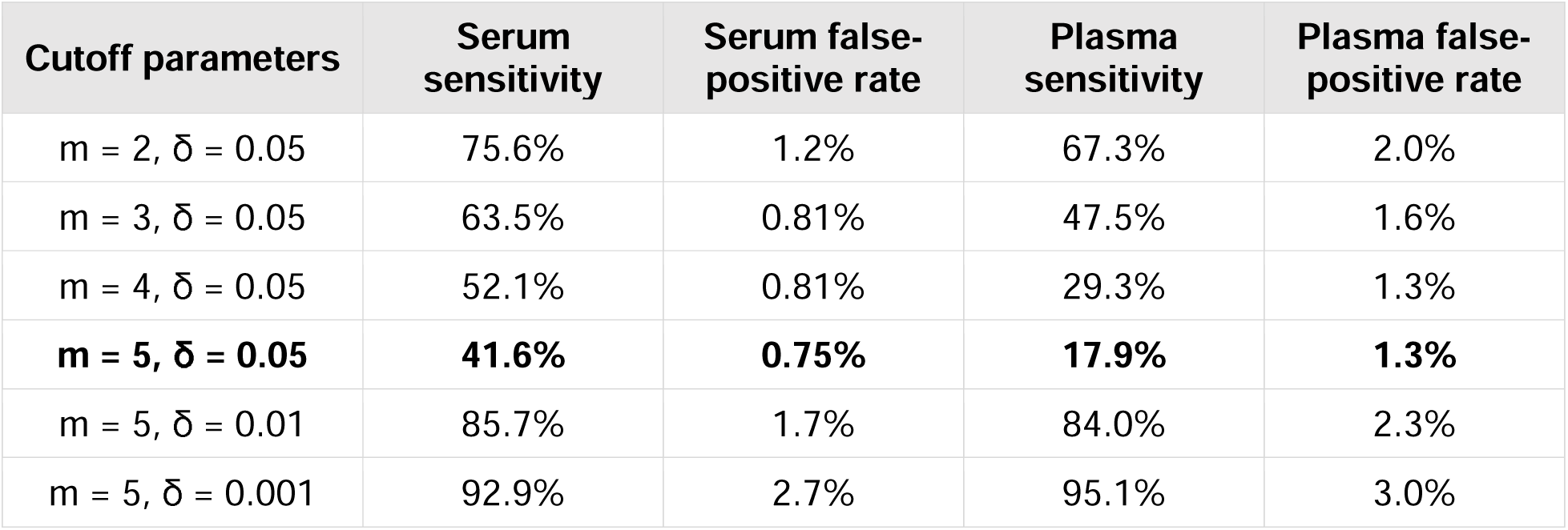
Mean cross-validated sensitivity and false-positive rate using the top 20 up- and downregulated metabolite features. Cutoff definitions and evaluation rounds are as in Table 1.

**Table 3B.** Mean cross-validated sensitivity and false-positive rate using the top 50 up- and downregulated metabolite features. Cutoff definitions and evaluation rounds are as in Table 1.

| Cutoff parameters | Serum sensitivity | Serum false-positive rate | Plasma sensitivity | Plasma false-positive rate |
| --- | --- | --- | --- | --- |
| $m = 2, \delta = 0.05$ | 96.8% | 1.0% | 91.7% | 1.8% |
| $m = 3, \delta = 0.05$ | 92.2% | 0.38% | 83.6% | 1.1% |
| $m = 4, \delta = 0.05$ | 86.9% | 0.0% | 72.1% | 0.80% |
| **$m = 5, \delta = 0.05$** | **79.5%** | **0.0%** | **59.0%** | **0.80%** |
| $m = 5, \delta = 0.01$ | 97.9% | 0.50% | 96.3% | 1.3% |
| $m = 5, \delta = 0.001$ | 98.7% | 0.80% | 97.9% | 1.6% |

**Table 3C.** Mean cross-validated sensitivity and false-positive rate using the top 100 up- and downregulated metabolite features. Cutoff definitions and evaluation rounds are as in Table 1.

| Cutoff parameters | Serum sensitivity | Serum false-positive rate | Plasma sensitivity | Plasma false-positive rate |
| --- | --- | --- | --- | --- |
| $m = 2, \delta = 0.05$ | 98.5% | 1.2% | 95.7% | 1.2% |
| $m = 3, \delta = 0.05$ | 97.0% | 0.50% | 90.7% | 1.1% |
| $m = 4, \delta = 0.05$ | 93.9% | 0.19% | 82.8% | 0.20% |
| **$m = 5, \delta = 0.05$** | **90.5%** | **0.19%** | **73.6%** | **0.20%** |
| $m = 5, \delta = 0.01$ | 98.3% | 0.63% | 97.3% | 0.50% |
| $m = 5, \delta = 0.001$ | 98.8% | 0.81% | 98.1% | 1.2% |

### Smaller panels of highly ranked features reduce sensitivity

At the matched setting m = 5 and δ = 0.05, the top-20, top-50, and top-100 analyses yielded serum sensitivities of 41.6%, 79.5%, and 90.5%, with false-positive rates of 0.75%, 0.0%, and 0.19%, respectively (Tables 3A–C). Corresponding plasma sensitivities were 17.9%, 59.0%, and 73.6%, with false-positive rates of 1.3%, 0.80%, and 0.20%. The full feature sets achieved sensitivities of 95.5% in serum and 90.3% in plasma, both with reported mean false-positive rates of 0.0%. At m = 4 and δ = 0.05, the top-100 analysis achieved sensitivities of 93.9% and 82.8%, with false-positive rates of 0.19% and 0.20%; the corresponding fractions of missed cancers were 6.1% and 17.2%. These comparisons favor broader feature sets under the reported analysis, but do not quantify individual contributions or establish statistical significance of performance differences. They are consistent with information distributed beyond the most strongly dysregulated features. Interpretation also depends on whether feature ranking and selection were confined to training data, and the small control groups limit precision at very low false-positive rates.

### Shared biochemical patterns across serum and plasma cohorts

We examined features with concordant changes across the cohorts to assess the biochemical organization of the OC-associated profile (Table 4). We identified 239 concordantly altered annotated features, listed in the supplementary workbook systemotype metabolites.xlsx. Features were grouped into biochemical classes because individual identities remain subject to uncertainty in untargeted metabolomics ^23^. Despite differences in sample matrix and independent processing, both cohorts showed alterations across similar biochemical classes. This concordance supports a shared pattern of association with OC, although class-level agreement alone does not establish coordinated regulation or exclude confounding.

**Table 4.**
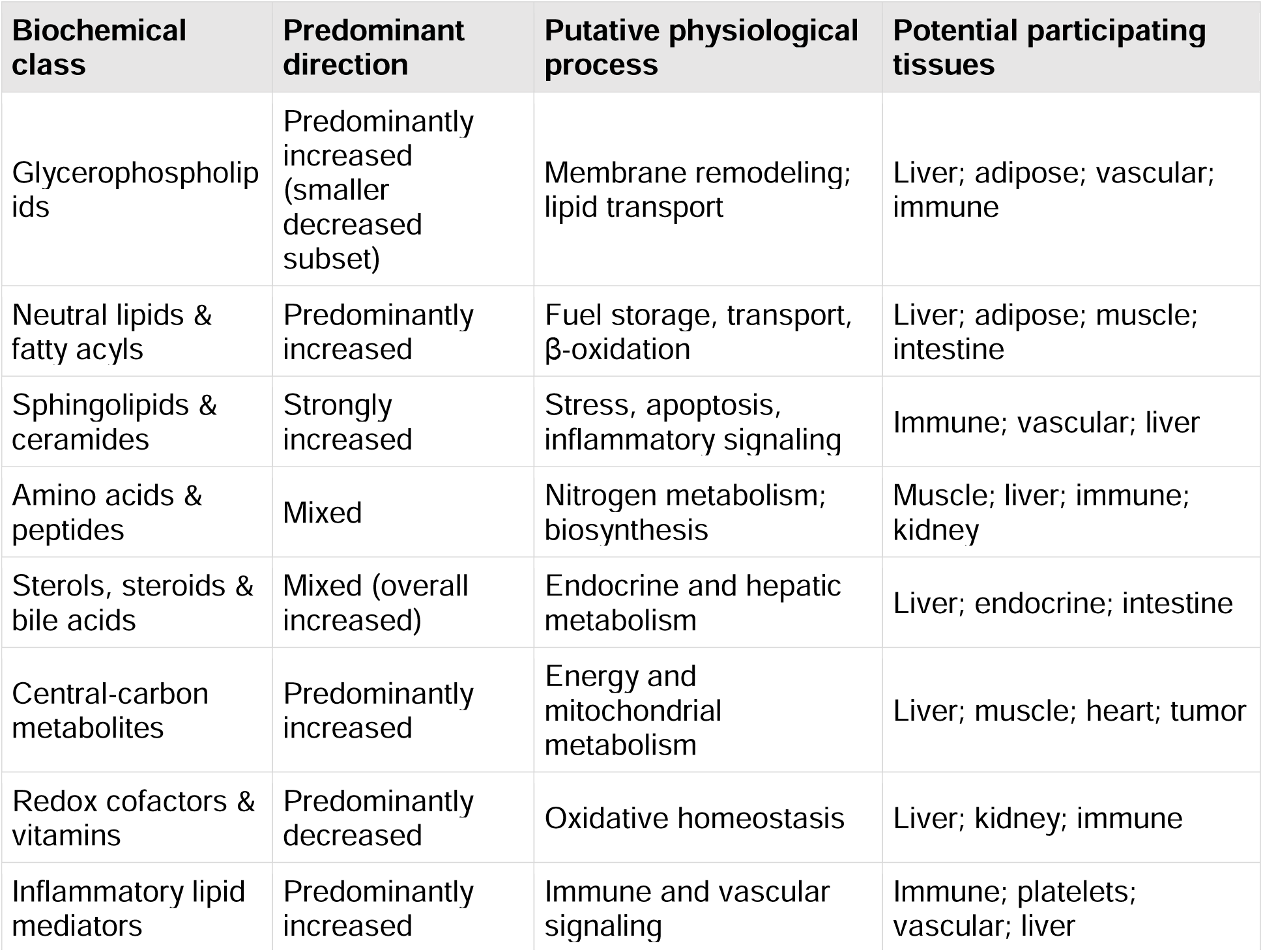
Biochemical classes represented among concordantly altered serum and plasma features. Physiological processes and participating tissues are proposed interpretations, not measured tissue contributions.

Lipid-associated features accounted for most concordant changes, including glycerophospholipids, neutral lipids, and sphingolipids. Alterations also involved amino-acid, central-carbon, steroid, and redox-associated metabolites. These classes participate in membrane remodeling, lipid transport, nutrient allocation, oxidative balance, endocrine regulation, and inflammatory signaling. Their breadth motivates an integrated physiological interpretation of the circulating profile, while the data themselves establish associations at the feature and class levels.

### Physiological interpretation of the shared metabolomic profile

The affected biochemical classes participate in processes maintained by multiple organs. Glycerophospholipids and neutral lipids are involved in hepatic, adipose, intestinal, and vascular metabolism ^24,25^. Sphingolipids contribute to membrane biology, stress responses, apoptosis, and immune signaling ^26^, while amino-acid metabolites reflect protein turnover and nutrient exchange among tissues ^27^. Sterol and bile-acid metabolism links intestinal, hepatic, and endocrine physiology; central-carbon and redox metabolites participate in energy metabolism and oxidative homeostasis. Circulating concentrations integrate production, consumption, transport, modification, and clearance ^4,5^. These associations identify plausible physiological contributors without assigning metabolites to specific tissues of origin.

### A proposed circulating Systemotype

As summarized in Table 4 and Figure 3, the shared profile spans lipid transport, lipoprotein remodeling, membrane turnover, and fuel allocation ^25^; sphingolipid-associated stress and immune signaling ^26^; and amino-acid and nitrogen metabolism ^24^. Steroid, bile-acid, central-carbon, and redox-associated features extend this profile to endocrine, hepatic, intestinal, and energy metabolism. The physiological interpretations overlap because these pathways are connected across tissues.

**Figure 3.**
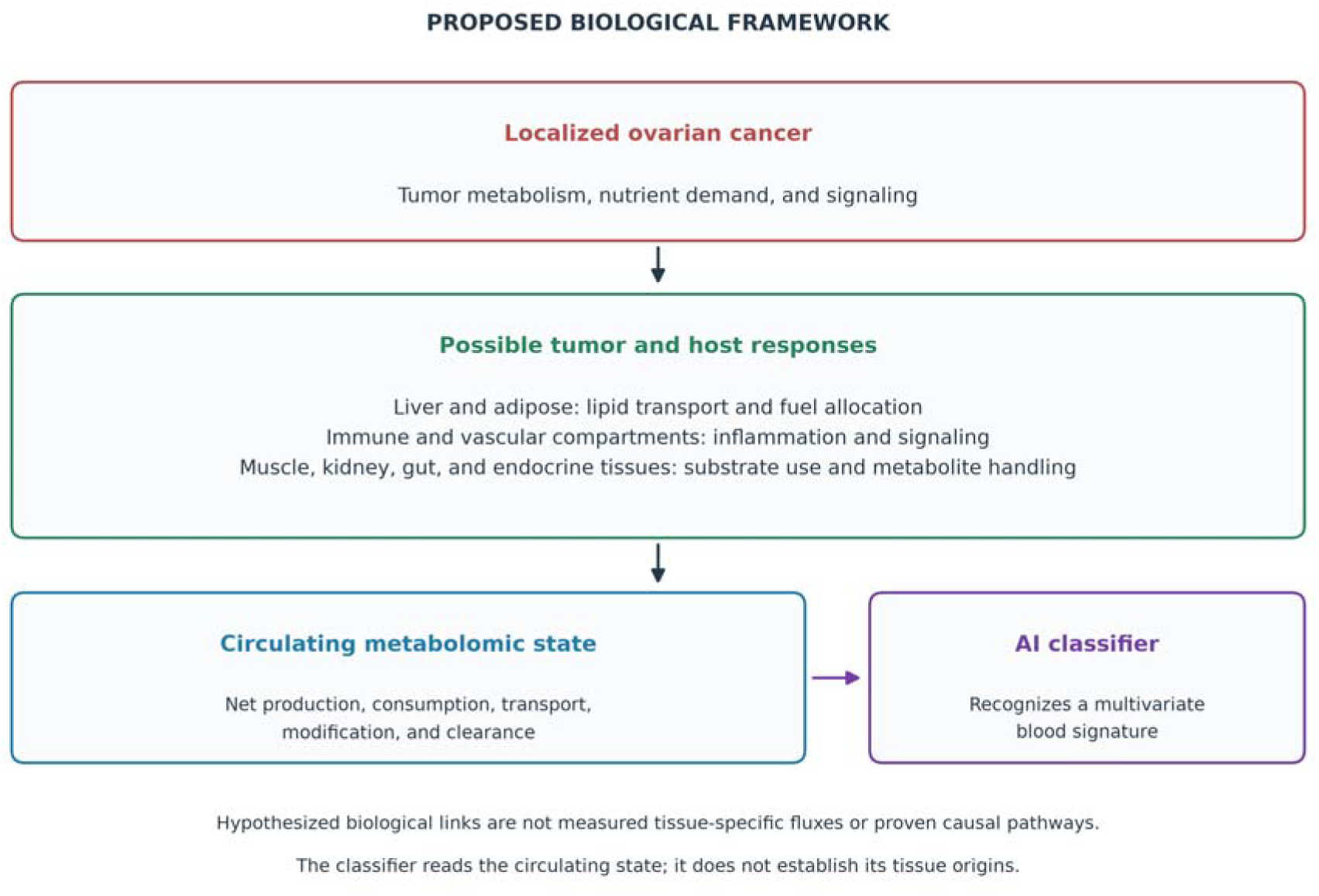
Proposed biological framework for a circulating Systemotype. Localized OC may perturb tumor and host metabolism, inflammatory and vascular signaling, nutrient demand, and communication among tissues. These responses could influence several physiological systems and alter the circulating metabolome ^24,28^. Organ-resolved fluxes and causal tissue contributions were not measured in this study; the model is a hypothesis for the observed multivariate blood profile.

Potential contributors include the liver, adipose tissue, immune and vascular compartments, skeletal muscle, kidneys, gastrointestinal tract, endocrine tissues, and tumor microenvironment ^24,29,30^. The net balance of metabolite production and handling across these compartments determines circulating concentrations ^4,5^. An altered blood profile can therefore be consistent with host responses without demonstrating dysfunction in each participating organ.

We propose that this distributed profile reflects a circulating Systemotype. In this model, a localized tumor perturbs processes integrated through communication among tissues. Adaptations in lipid transport, nutrient processing, fuel mobilization, immune signaling, and metabolite handling could amplify the consequences of a small lesion into a measurable circulating phenotype ^10,28^. This is a mechanistic hypothesis prompted by the metabolomic associations, rather than a mechanism established by the classifier.

### Published evidence and boundaries of the Systemotype hypothesis

Published studies provide context for possible effects of OC beyond the ovary. Metabolomic differences have been reported between early-stage OC and benign pelvic or ovarian masses ^31,32,33^, but our cohorts compare OC with healthy controls and do not test this distinction. The literature discussed below supports possible contributions from skeletal muscle, immune and inflammatory processes, and systemic metabolism. Evidence for early intrinsic renal dysfunction, hepatic synthetic impairment, platelet responses, and steroid-axis changes is more limited or indirect. Absence of evidence for organ failure does not exclude subtler changes in metabolite exchange.

### Skeletal muscle

A retrospective human study associated sarcopenia at stage I/II OC diagnosis with poorer survival ^34^. This observation does not establish that the tumor caused the muscle deficit. In an orthotopic, immunocompetent ID8 mouse model, muscle weakness and mitochondrial stress developed before severe metastasis and muscle atrophy, with impaired pyruvate oxidation ^35^. The investigators could not exclude all early metastatic spread. A subsequent study found that mitochondrial-targeted SkQ1 treatment partially prevented weakness without preventing atrophy or improving survival ^36^. These experiments support a contribution of mitochondrial dysfunction to muscle weakness during OC progression, but do not directly connect muscle metabolism to the circulating features measured here.

### Systemic immune and inflammatory responses

Studies of stage I OC and broader ovarian-neoplasm populations associate systemic inflammatory indices with disease status or prognosis ^37,38,39^. These indices indicate changes in circulating cell populations but do not establish a tumor-specific mechanism. Enhanced tryptophan degradation in OC has also been associated with soluble immune-activation markers ^40^. Work on tumor immunity and indoleamine 2,3-dioxygenase (IDO) provides a possible mechanistic context ^41,42,43,44^. Together, these observations support immune-associated contributions to circulating metabolism, although they do not establish that the tryptophan signal originates exclusively from immune cells or is specific to early-stage disease.

### Systemic intermediary metabolism

Metabolomic studies report alterations in circulating fuel and amino-acid metabolism in OC ^32^, and a lipidomic study included symptomatic controls with benign conditions ^45^. Other studies identify changes in amino-acid, phospholipid, and fatty-acid metabolism ^46,47,48^, with time-dependent remodeling also reported in a high-grade serous OC mouse model ^49^. Associations between tryptophan degradation and immune activation caution against assigning the circulating tryptophan signal solely to hepatic metabolism ^40^. Bile-acid pathways may participate in cancer biology ^50^, but the evidence cited here does not establish hepatic synthetic failure in early-stage OC. Such failure is not a necessary prediction of the Systemotype model.

### Renal function and circulating markers

The cited renal-marker studies do not establish intrinsic kidney dysfunction caused by early-stage OC. Cystatin C can be influenced by tumor-related factors ^51^, while HE4 rises with renal impairment independently of OC ^52,53^. NGAL/lipocalin-2 can reflect tumor biology as well as processes relevant to kidney injury ^54,55^. In treated patients, olaparib can increase serum creatinine through effects on renal transport without a corresponding reduction in measured filtration ^56^. These findings emphasize confounding and clearance as considerations when interpreting circulating metabolites, rather than proving early renal involvement in the observed profile.

### Vascular and platelet responses

Lysophosphatidic acid (LPA) illustrates the difficulty of separating biological effects from pre-analytical influences. Differences among healthy, benign, and malignant groups have been reported ^57^, but overlap and ex vivo generation complicate interpretation ^58^. Tumor-associated IL-6 signaling and hepatic thrombopoietin production provide a mechanism for paraneoplastic thrombocytosis in OC ^59^. This mechanism is relevant to tumor–host communication, but the cited evidence does not establish that platelet responses explain the metabolomic profile of early-stage cases in this study.

### Steroid metabolism

Evidence for altered steroid metabolism in early-stage OC remains indirect in the studies considered here. Postmenopausal ovarian steroid production provides physiological context ^60^, and an endometrial-cancer metabolomic study demonstrates the potential diagnostic information in steroid-associated profiles ^61^. Neither observation establishes an OC-specific steroid signature or independence from inflammatory effects. Targeted steroid measurements in early OC and appropriately matched benign controls would test this proposed component of the Systemotype.

### Interpreting a distributed blood signal

A central question is how an anatomically localized tumor could be associated with a detectable signal in peripheral blood. The 239 concordantly altered annotated features span several biochemical classes rather than a single pathway. This breadth is compatible with a combination of tumor-derived metabolites and host responses. It does not, by itself, distinguish their relative contributions or demonstrate an emergent state.

The affected classes participate in lipid transport, membrane remodeling, inflammatory signaling, oxidative homeostasis, nutrient allocation, and endocrine and energy metabolism. These processes operate in both tumor and host tissues ^10,62^. The Systemotype framework offers one interpretation of their joint appearance in blood; tissue-resolved or longitudinal measurements are needed to test it.

### Testing the link between tumor burden and systemic responses

The literature supports investigating muscle metabolism, inflammatory responses, and systemic nutrient handling as contributors to the circulating profile. It provides weaker support for assigning the observed changes to early intrinsic renal dysfunction, hepatic synthetic impairment, or a specific steroid or platelet mechanism. Physiological participation should therefore be distinguished from organ injury or failure.

Most human evidence is cross-sectional. Mouse experiments support temporal and mechanistic links between OC progression and muscle dysfunction, while retaining limitations concerning early metastatic spread ^35,36^. Human studies associate greater disease burden and treatment selection with baseline sarcopenia ^63^. Changes during neoadjuvant chemotherapy are difficult to interpret because treatment and tumor reduction occur together ^63,64,65^. A prospective study with repeated metabolomic measurements before and after tumor removal, appropriately timed to separate postoperative responses from recovery, could test whether a circulating signature tracks disease burden.

## Discussion

### The Systemotype as a testable physiological framework

We define a Systemotype as an integrated physiological state arising from interactions among organs, tissues, cells, and molecular networks and reflected in measurable molecular phenotypes. Disease may alter metabolic demand, inflammatory signaling, immune activity, and nutrient allocation, changing metabolite production and handling across tissues ^8,9,10^. The present findings motivate this framework but do not demonstrate that each observed feature is part of a coordinated causal response.

### Implications for metabolomic classification

A distributed physiological response could help explain why broad metabolomic profiles contain classification information beyond a small set of strongly altered features ^9,11^. Features arising from different processes may provide complementary information. However, predictive performance alone does not identify those processes, establish interactions among them, or rule out demographic and pre-analytical confounding.

The results provide a retrospective proof of concept for classifying OC within two separately modeled cohorts. They also motivate a framework for testing how localized disease is reflected in circulating metabolism. Clinical utility and biological mechanism remain separate questions requiring prospective validation and targeted experiments, respectively.

The combination of class-level concordance and reduced sensitivity in smaller panels is consistent with a distributed metabolomic association. It supports further study of the Systemotype hypothesis, without establishing its causal basis or the suitability of the current models for screening.

### Limitations and next steps

The cohorts are small and retrospective, with only 32 serum controls and 20 plasma controls. Repeatedly partitioning these samples does not increase the number of independent participants or establish precise population estimates at very high specificity. The plasma model was developed separately and is not external validation of the serum model. Performance at retrospectively compared operating points requires confirmation using prespecified thresholds and locked models in independent, prospectively collected populations ^17,18,19,20^. All label-informed feature ranking, selection, and threshold estimation must be confined to training data within each outer fold; otherwise, performance may be optimistic. The healthy-control design does not establish specificity against benign gynecologic disease, other cancers, or inflammatory and metabolic conditions. Future studies should assess demographic and pre-analytical confounding, report participant-level uncertainty and calibration, and evaluate predictive values at the intended-use prevalence. Longitudinal and tissue-resolved measurements are needed to test the proposed biological interpretation.

Blood metabolomic profiles distinguished OC from healthy controls under the reported within-cohort evaluation and showed shared biochemical patterns across serum and plasma. These observations support investigation of a circulating Systemotype as a physiological framework for metabolomic classification. Prospective validation and mechanistic studies are required before extending this interpretation to clinical diagnosis or population screening.

## Data and code availability

Processed metabolite intensity matrices, annotation tables, and training and cross-validation code will be made available by the authors under appropriate data-use terms. Compound annotations were interpreted in the context of the Metabolomics Standards Initiative (MSI) reporting framework ^23^, with annotation probabilities generated using Panome Bio’s MassID workflow ^66^. The Systemotype code is released under the MIT License and is available at 10.5281/zenodo.22679113.

## Author contributions

JS conceived the Systemotype framework, performed the pathway and organ-system analyses, and developed the overall methodology. HZ developed the AI analysis tools and blood-diagnostic software. JLC oversaw the experimental work, and BB provided medical guidance. JS drafted the manuscript. All authors reviewed and revised it.

## Data Availability

https://github.com/hzhou3ga/Systemotype

## Acknowledgements

We thank Jessica Forness for her assistance in proofreading and manuscript preparation.

## Methods

### Study samples

Two independent cohorts of human serum and EDTA-plasma samples from patients with OC and healthy donors were obtained through certified sources and submitted to Panome Bio for untargeted metabolomic analysis. The serum cohort contained 59 OC samples and 32 healthy controls; the plasma cohort contained 63 OC samples and 20 healthy controls.

### Sample preparation

A 50 µL aliquot of each serum or plasma sample was transferred to a solid-phase extraction (SPE) system. A separate 10 µL aliquot from each sample was pooled to create a quality-control (QC) sample. Each well received 200 µL of 1:1 acetonitrile:methanol (ACN:MeOH), was shaken for 1 min at room temperature at 360 rpm, and was incubated for 10 min. Next, 150 µL of 2:2:1 MeOH:ACN:water was added, followed by another 10 min extraction step. Polar metabolites were eluted into a 96-well collection plate using a positive-pressure manifold. Elution was repeated with 100 µL of 2:2:1 MeOH:ACN:water into the same plate. Polar eluates were covered and stored at −80 °C until liquid chromatography–mass spectrometry (LC–MS) analysis. Lipids were then eluted from the SPE plates into a new collection plate by two washes with 500 µL of 1:1 methyl tert-butyl ether (MTBE):MeOH using the positive-pressure manifold. Combined lipid eluates were dried under nitrogen at room temperature and reconstituted in 200 µL of 1:1 isopropanol (IPA):MeOH before LC–MS analysis.

### Metabolomic assays

#### LC MS analysis of polar metabolites

Mobile phase A contained 20 mM ammonium bicarbonate, 0.1% ammonium hydroxide, 5% ACN, and 2.5 mM medronic acid; mobile phase B was 95% ACN. A 2 µL aliquot of polar extract was analyzed by hydrophilic interaction liquid chromatography–mass spectrometry (HILIC–MS) at 250 µL/min using the following gradient: 0–1 min, 90% B; 1–12 min, 90–35% B; 12–12.5 min, 35–20% B; and 12.5–14.5 min, 20% B. The column was re-equilibrated with 20 column volumes of 90% B. Mass spectra were acquired over the reported mass range of 67–1500 Da at one scan/s and a resolving power of 120,000 in positive and negative ionization modes. Tandem mass spectrometry (MS/MS) data were acquired iteratively in a data-dependent manner with a 1.3 m/z isolation window.

#### LC MS analysis of lipids

Mobile phase A contained 5:3:2 water:ACN:IPA, 10 mM ammonium formate, and 5 µM Agilent deactivator additive; mobile phase B contained 1:9:90 water:ACN:IPA and 10 mM ammonium formate. A 4 µL aliquot of lipid extract was analyzed by reversed-phase LC–MS on a Waters Acquity Premier HSS T3 column (2.1 × 100 mm) at 400 µL/min. The gradient was: 0–2.5 min, 15–50% B; 2.5–2.6 min, 50–57% B; 2.6–9 min, 57–70% B; 9–9.1 min, 70–93% B; 9.1–11 min, 93–96% B; 11–11.1 min, 96–100% B; 11.1–12 min, 100% B; and 12–12.2 min, 100–15% B. The column was re-equilibrated for 3.8 min. Mass spectra were acquired over m/z 100–1700 at one scan/s in positive and negative ionization modes. MS/MS data were acquired iteratively in a data-dependent manner with a 1.3 m/z isolation window.

## Data processing

### Metabolite detection and identification

Metabolite signals (features) were detected using in-house LC–MS peak-detection and curation software and aligned across samples. The reported contamination filter removed features with intensities greater than one-third of the corresponding intensity in the QC sample. Redundant signals, including isotopes, adducts, and fragments, were identified by clustering and metabolite assignment using in-house software. Features were annotated against a database of metabolites drawn from RefMet, LIPID MAPS, and HMDB using isotope patterns and MS/MS fragmentation data when available. Annotation confidence was interpreted in the context of the MSI framework ^23^, and identification probabilities were generated using MassID ^66^.

### Metabolite annotation confidence

The annotation workflow used four reported confidence categories. Level 1 assignments were supported by retention times and MS/MS spectra from authentic standards and an isotope-pattern match. Level 2 assignments required an isotope-pattern similarity greater than 90% (reverse dot product), an MS/MS entropy similarity greater than 50%, and a predicted retention-time difference of less than 2 min. Level 3 assignments required isotope-pattern and predicted retention-time matches using the same thresholds. Level 4 features had no matching compound in the metabolite database. These operational criteria describe the annotation workflow; predicted retention times do not substitute for authentic-standard confirmation. For lipids, the best-matching species was reported, but the LC–MS methods did not resolve acyl-chain double-bond positions. Positional annotations should therefore not be interpreted as experimentally established.

### Data normalization and curation

Data from the four assays were concatenated. Features were excluded if the coefficient of variation across QC samples exceeded 25%. Missing values were imputed with half the minimum detected intensity for the corresponding feature. Intensities were log2-transformed before statistical analysis. Annotations and intensities were manually reviewed for concordance and accuracy.

### Summary of experimentally identified metabolite features

Batch 1 comprised 3,433 metabolite features from 91 serum samples (59 OC, 32 healthy controls). Batch 2 comprised 3,625 features from 83 plasma samples (63 OC, 20 healthy controls). Each sample’s features were partitioned into lipid_pos, lipid_neg, polar_pos, and polar_neg assay sets. Annotations followed standard lipid and small-molecule nomenclature, with lipid classes assigned from the leading abbreviation. Small-molecule annotations were interpreted using reference metabolome resources ^67^. Figure 1 summarizes the workflow.

### Boosted decision tree classifier

Samples were classified as OC or healthy control using boosted-tree regression ^21^. Gradient boosting fits an additive sequence of trees to improve the prediction objective ^21,68^. Each tree had a maximum depth of six. The ensemble score was

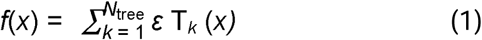

where T is the kth decision tree, ε is the learning rate, and Ntree is the number of trees. We used ε = 0.05 and Ntree = 1,000; these values were chosen empirically and were not optimized. Each metabolite intensity contributed one component of the feature vector. Assay-level feature counts were 2,364 (lipid_pos), 253 (lipid_neg), 302 (polar_pos), and 514 (polar_neg) in serum, and 1,160, 570, 668, and 1,227, respectively, in plasma. Regression targets were 1 for OC and 0 for healthy controls. Each assay was modeled separately. A sample was called positive when its score exceeded a training-derived cutoff. Assay-level calls were combined using an OR rule: any positive assay classified the sample as OC; all assays had to be negative for a healthy-control classification.

### Fivefold cross validation and cutoff determination

Samples were partitioned into five groups. Within each round of the 50 tests, randomization consisted of 1,000,000 swaps of the group assignments of two randomly selected samples. Each group then served once as the held-out test set, while the remaining four groups formed the training set. Test labels were excluded from model training and threshold estimation. Leave-one-out cross-validation within the training set provided scores for deriving the decision threshold. This procedure was repeated for 50 rounds with different pseudorandom seeds, yielding 250 held-out evaluations per cohort. Reported performance values are means across these evaluations. This procedure randomizes held-out fold assignment; it is not a label-permutation test of a null hypothesis.

To determine the cutoff, internal cross-validation scores from the training set were sorted in descending order. Let N be the number of OC samples ranked above the highest-scoring healthy control. The cutoff was

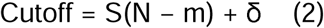

where S(N − m) is the score of the OC sample at rank N − m, m is a nonnegative integer specifying the ranking gap, and δ is an additive offset. Thus, m = 0 selects the OC sample immediately above the highest-scoring healthy control, provided such a sample exists. The stated rule requires N > m. We report several (m, δ) settings to describe the observed performance trade-off. An intended-use operating point must be prespecified before independent evaluation.

### Secondary analysis of dysregulated annotations

For biological interpretation, features were categorized as increased or decreased in OC within each cohort using the Mann–Whitney U test criterion |z| > 2 ^22^. The resulting tables contained 1,354 increased and 806 decreased features in serum, and 1,197 increased and 840 decreased features in plasma. Cross-cohort concordance required matching analytical assays, exact annotation labels, and the same direction of change. Because annotations can contain isomeric or multiple-candidate identities, concordance represents annotation-level rather than definitive compound-level replication. Lipid classes were assigned from the leading abbreviation, such as Cer, PC, PE, TG, DG, or LPC. Within each cohort, class enrichment among increased versus decreased features was assessed with a two-sided Fisher exact test ^69^. Six lipid classes were tested per cohort, so class-level P values require consideration of multiple testing ^70^. This biological analysis was descriptive. It is distinct from the restricted-feature classification analyses in Tables 2 and 3, which require feature selection to be nested within the training folds.

